# Viral kinetics of sequential SARS-CoV-2 infections

**DOI:** 10.1101/2023.03.03.23286775

**Authors:** Stephen M Kissler, James A Hay, Joseph R Fauver, Christina Mack, Caroline G Tai, Deverick J Anderson, David D Ho, Nathan D Grubaugh, Yonatan H Grad

## Abstract

The impact of a prior SARS-CoV-2 infection on the progression of subsequent infections has been unclear. Using a convenience sample of 94,812 longitudinal RT-qPCR measurements from anterior nares and oropharyngeal swabs, we compared the SARS-CoV-2 viral kinetics of first *vs*. second infections, adjusting for viral variant, vaccination status, and age. Relative to first infections, second infections usually featured a lower peak viral concentration and faster clearance time, especially in individuals who received a vaccine dose between their first and second infection. Furthermore, a person’s relative (rank-order) viral clearance time, compared to others infected with the same variant, was similar across first and second infections; that is, individuals who had a relatively fast clearance time in their first infection tended to also have a relatively fast clearance time in their second infection. These findings provide evidence that, like vaccination, immunity from a prior SARS-CoV-2 infection shortens the duration of subsequent acute SARS-CoV-2 infections principally by reducing viral clearance time. Additionally, there appears to be an inherent element of the immune response, or some other host factor, that shapes a person’s relative ability to clear SARS-CoV-2 infection that persists across sequential infections.

## Introduction

An estimated 65% of the US population had at least two SARS-CoV-2 infections by November 2022, but the impact of prior infection on disease course in subsequent infections has been debated.^1^ Some evidence indicates SARS-CoV-2 infection provides a temporary reduction in reinfection risk^2^ and a durable reduction in the risk of COVID-19-related hospitalization and death,^3^ while a handful of studies suggest that an initial SARS-CoV-2 infection may limit recovery from COVID-19 infections.^4^ These contrasting findings may result from biases that can arise in population-level studies when differences in exposure history, vaccination status, and comorbidities are not fully accounted for. Controlling for such factors is a major challenge given geographically and temporally heterogeneous interventions, whereas examining the dynamics of SARS-CoV-2 infections at the individual level can facilitate adjusting for these biases.

Reverse transcription quantitative PCR (RT-qPCR) conducted from clinical samples collected at multiple time points offer an objective, quantitative metric of SARS-CoV-2 kinetics and can inform on key aspects of immune response and clinical progress. Such data have been used to specify how vaccination history, antibody titer and viral lineage together shape SARS-CoV-2 proliferation and clearance during an acute infection,^5^ which in turn can inform the clinical management of COVID-19^6^ and help interpret epidemiological trends.^7^ Viral kinetics therefore offer a promising metric for clarifying the impact of an initial SARS-CoV-2 infection on subsequent infections and for translating those findings into medical and public health guidance.

The impact of vaccination and variant on SARS-CoV-2 viral kinetics have been well described elsewhere.^8–13^ Infections with Delta lineages feature a higher peak viral concentration than Alpha or Omicron infections, and vaccination speeds up the clearance of SARS-CoV-2 across lineages.^11^ However, the impact of SARS-CoV-2 infection-conferred immunity on peak viral concentration, viral proliferation, and viral clearance in subsequent infections is less well characterized. Furthermore, it has been unclear to what extent attributes of SARS-CoV-2 kinetics, such as peak viral load or clearance rate, persist across an individual’s successive infections.

Here, we collected and analyzed 94,812 SARS-CoV-2 RT-qPCR viral concentration measurements taken from longitudinal clinical samples in players, staff, and affiliates of the National Basketball Association (NBA) between March 11^th^, 2020, and July 28^th^, 2022. For the subset of individuals who were infected twice during the study period, we measured changes in viral kinetics between first and second infections and determined the extent to which viral kinetic features persisted across infections.

## Methods

### Study design

Between March 11^th^, 2020, and July 28^th^, 2022, the NBA conducted regular surveillance for SARS-CoV-2 infection among players, staff, and affiliates as part of an occupational health program. This included frequent viral testing (often daily during high community COVID-19 prevalence) using a variety of platforms, but primarily via nucleic acid amplification tests, as well as clinical assessment including case diagnosis and symptom tracking. To assess viral concentration, RT-qPCR tests were conducted when possible, using anterior nares and oropharyngeal swabs collected by a trained professional and combined into a single viral transport media. Cycle threshold (Ct) values were obtained from the Roche cobas target 1 assay. Ct values were converted to genome equivalents per milliliter using a standard curve.^14^ Data on participant age and vaccination status were collected where possible. Viral lineages were assigned using whole-genome sequencing (**Supplementary Methods**), when feasible. This resulted in a longitudinal dataset of 424,401 SARS-CoV-2 tests with clinical COVID-19 history and demographic information for 3,021 individuals.

During the data collection period, 3,346 infections were identified among the 3,021 individuals. These infections reflected the timing, intensity, and lineage composition of SARS-CoV-2 transmission in the broader United States (**Figure 1**). Of these infections, we identified 1,989 “well-documented” infections that were sufficiently sampled to infer viral kinetics (**Table 1**), as defined by at least one RT-qPCR test with cycle threshold (Ct) value under 32 and three tests with Ct values under 40^11^ One individual had four total infections, and we omitted their third and fourth from the analysis. Of the well-documented infections, 193 were second infections, as evidenced by a previous positive test more than 30 days prior to infection or a clinical history of prior infection (**Figure 1, Supplementary Table 1**).

**Table 1.** Characteristics of the well-documented infections. Counts by lineage, vaccination status, age group, and cardinality of infection (first, second, third, or fourth) for the 1,989 well-documented infections. Well-documented infections are those with at least one RT-qPCR Ct < 32 and three Ct < 40 (the limit of detection). Infections occurred between 11^th^ March 2020 and 28^th^ July 2022.

| Statistic | Category | N | % |
| --- | --- | --- | --- |
| **Total** |  | **1989** | **100** |
| **Lineage +**<br>**vaccination status at**<br>**time of infection** | **Alpha** | **49** | **2.5** |
|  | Unvaccinated | 24 | 1.2 |
|  | Vaccinated | 4 | 0.2 |
|  | Unknown primary vaccination status | 21 | 1.1 |
|  | Unboosted | 22 | 1.1 |
|  | Boosted | 0 | 0.0 |
|  | Unknown booster status | 27 | 1.4 |
|  | **Delta** | **191** | **9.6** |
|  | Unvaccinated | 7 | 0.4 |
|  | Vaccinated | 123 | 6.2 |
|  | Unknown primary vaccination status | 61 | 3.1 |
|  | Unboosted | 99 | 5.0 |
|  | Boosted | 10 | 0.5 |
|  | Unknown booster status | 82 | 4.1 |
|  | **BA.1/BA.2** | **1400** | **70.4** |
|  | Unvaccinated | 6 | 0.3 |
|  | Vaccinated | 994 | 50.0 |
|  | Unknown primary vaccination status | 400 | 20.1 |
|  | Unboosted | 117 | 5.9 |
|  | Boosted | 778 | 39.1 |
|  | Unknown booster status | 507 | 25.5 |
|  | **BA.4/BA.5** | **71** | **3.6** |
|  | Unvaccinated | 0 | 0.0 |
|  | Vaccinated | 53 | 2.7 |
|  | Unknown primary vaccination status | 18 | 0.9 |
|  | Unboosted | 0 | 0.0 |
|  | Boosted | 50 | 2.5 |
|  | Unknown booster status | 21 | 1.1 |
|  | **Other/Unspecified** | **278** | **14.0** |
|  | Unvaccinated | 98 | 4.9 |
|  | Vaccinated | 81 | 4.1 |
|  | Unknown primary vaccination status | 99 | 5.0 |
|  | Unboosted | 92 | 4.6 |
|  | Boosted | 55 | 2.8 |
|  | Unknown booster status | 131 | 6.6 |
| **Age group** | 0-29 | 816 | 41.0 |
|  | 30-49 | 876 | 44.0 |
|  | 50-99 | 295 | 14.8 |
|  | Not Reported | 2 | 0.1 |
| **Infection number** | 1 | 1796 | 90.3 |
|  | 2 | 193 | 9.7 |

**Figure 1.**
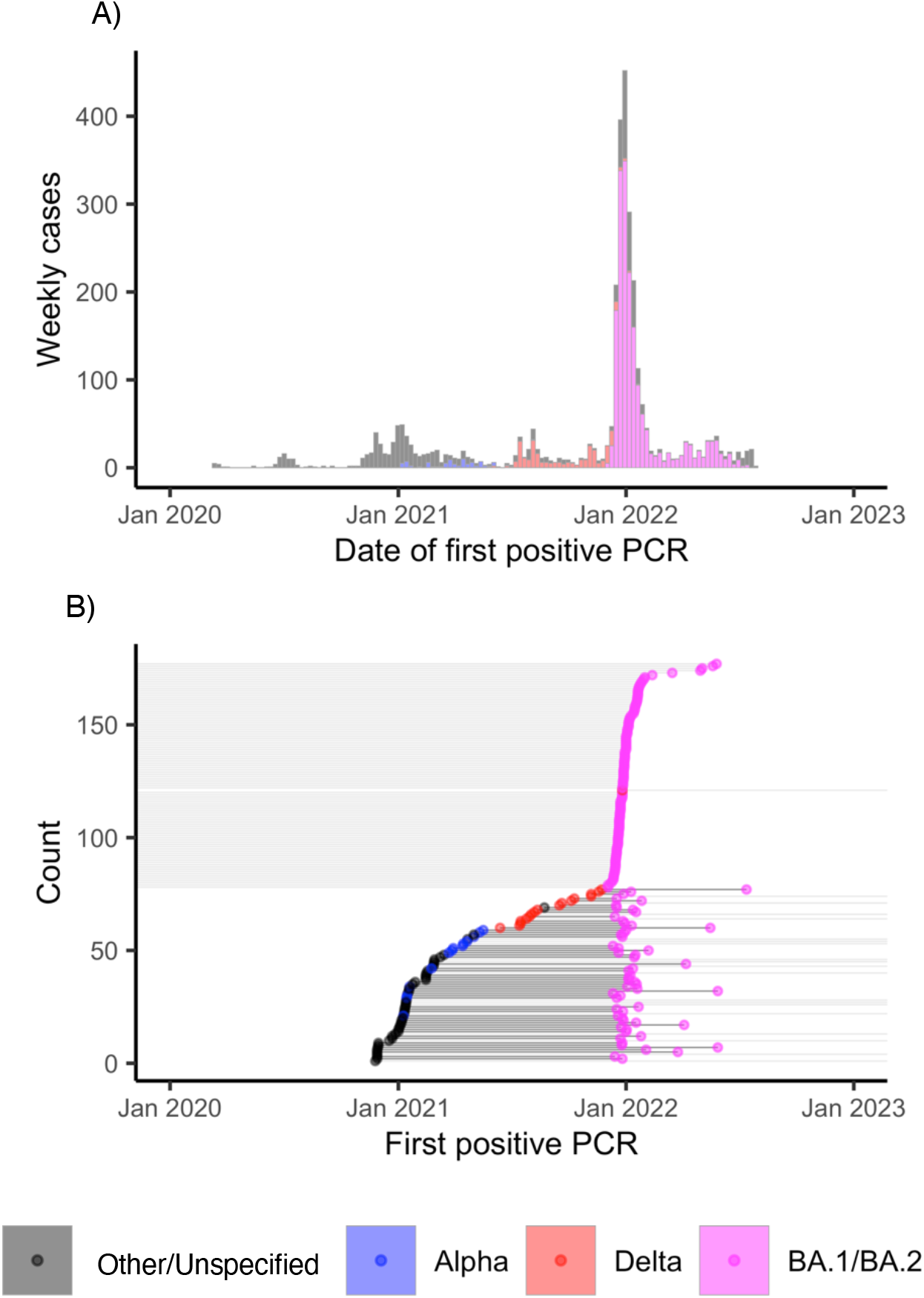
Onset times of repeat and overall infections in the dataset. (A): Histogram of first positive test dates for all recorded infections in full dataset (*n =* 3,346). Colors in both panels correspond to the SARS-CoV-2 variant category (Other/Unspecified: Black; Alpha: Blue; Delta: Red; BA.1/BA.2: Magenta), where Other/Unspecified include all non-Alpha, Delta, and Omicron lineages and any samples that could not be sequenced. (B): Date of the first positive test (points) for well-documented infections (*n =* 235) in individuals with an initial Alpha, Delta, or Other/Unspecified infection (n = 78 well-documented infections) and a second BA.1/BA.2 infection (n = 157 well-documented infections; see also **Supplementary Table 1, group 2**). Horizontal lines connect the points that correspond to multiple infections that belong to a single person. Lines that extend off the plot range to the left indicate the absence of a well-documented (though clinically confirmed) first infection, and lines that extend off the plot range to the right indicate the absence of a well-documented (though clinically confirmed) second infection.

Vaccination status (unvaccinated, fully vaccinated, or boosted) was assigned at the time of the first positive test for each infection. Full vaccination corresponded to 14 days following either the second dose of a Pfizer or Moderna vaccine or the first dose of a Johnson and Johnson/Janssen vaccine. A person was considered “boosted” 14 days after an additional Pfizer or Moderna dose following their initial vaccination course.

### Estimating viral kinetic parameters

We characterized the viral kinetics of the well-documented infections by fitting a hierarchical piecewise linear model to the viral concentration measurements on a logarithmic scale, following previous methods.^11,14^ The model captures the viral proliferation time (*i*.*e*., time from first possible detection to peak), peak viral concentration, and clearance time (*i*.*e*., time from peak to last possible detection) of acute SARS-CoV-2 infections. The hierarchical structure allowed us to estimate population-level differences in viral kinetic parameters associated with vaccination status, viral lineage, and history of a prior SARS-CoV-2 infection. We adjusted the model fits by the age group of the infected individual (0–29 years, 30–49 years, and 50+ years). Model parameters were fit using a Hamilton Monte Carlo algorithm implemented in R (version 4.1.2) and Stan (version 2.21.3).

### Study outcomes

We assessed whether Omicron BA.1/BA.2 viral kinetics differed according to whether the infection was the individual’s first SARS-CoV-2 infection (*n* = 1,241 well-documented infections) or their second infection (*n* = 159 well-documented infections; **Supplementary Table 1, group 1**), stratifying by booster status and adjusting for age. Then, for the subset of individuals with a first infection by a non-Omicron lineage (verified by whole-genome sequencing or an infection onset prior to November 11^th^, 2021) and a second infection with BA.1 or BA.2 (*n* = 235 well-documented infections among 177 individuals; **Supplementary Table 1, group 2**), we estimated (a) the difference in mean viral kinetics (proliferation time, clearance time, and peak viral load) between first and second infections and (b) the difference in viral kinetics of a second BA.1/BA.2 infection associated with the variant category of the first infection (Alpha, Delta, or other lineages). We note that individuals who underwent two infections may differ from the rest of the population in terms of their exposure patterns, immunity, and/or sampling patterns. Last, using the subset of individuals (*n* = 58; **Supplementary Table 1, group 3**) with two well-documented infections, we assessed whether a person’s relative individual-level viral kinetic features (proliferation time, clearance time, or peak viral load) persisted across infections.

### Statistical approach

We assessed differences in viral kinetic parameters across category subsets (*e*.*g*., by presence/absence of a prior infection or by vaccination status) by subtracting the relevant posterior draws and measuring the posterior probability mass of these differences that sat above/below 0, depending on the scenario. When fewer than 5% of these differenced posterior draws sat above/below zero, we took this as evidence of a significant difference.

To assess relative persistence in individual-level viral kinetic attributes across infections, we measured the Spearman correlation between the rank of the viral kinetic parameter in the first infection and the rank of the same parameter for the same person in the second infection, relative to all other infections caused by the same variant in individuals with two well-documented infections (**Supplementary Methods**). We used Spearman correlation because, as a rank-based measure, it is less affected by outliers and non-linear relationships between the viral kinetics in the first and second infection. As a sensitivity analysis, we repeated the analysis using the more common Pearson correlation (**Supplementary Methods**). To assess the statistical significance of the correlations, we conducted permutation tests, randomly assigning first- and second-infection viral kinetic parameters to different individuals and re-calculating the correlation. We then compared the true correlation to the distribution of correlations achieved through 100,000 permutations and concluded a significant difference when the true correlation was higher than 95% of the correlations obtained from permutation.

### Study oversight

This work was approved as “research not involving human subjects” by the Yale Institutional Review Board (HIC protocol # 2000028599), as it involved de-identified samples. This work was also designated as “exempt” by the Harvard Institutional Review Board (IRB20-1407).

## Results

### In BA.1/BA.2 infections, evidence of a previous infection correlates with faster clearance

Omicron BA.1/BA.2 infections in individuals with a history of prior infection (*n* = 159) featured a faster mean clearance time (4.9 days (4.5, 5.3)) than Omicron BA.1/BA.2 infections in individuals with no prior infection history (*n* = 1,241; 7.2 days (6.8, 7.5)) (**Figure 2A–B; Supplementary Table 1, group 1**). Even in individuals who had received a booster vaccine dose, BA.1/BA.2 clearance times were faster in individuals with a history of prior infection (5.1 days (4.5, 5.7)) than in those with no such history (7.1 days (6.7, 7.6); **Supplementary Table 2**). Omicron BA.1/BA.2 clearance rates were also faster in vaccine-boosted individuals with a history of prior infection (2.6 Ct/day (2.2, 3.0)) than in those without (2.0 Ct/day (1.8, 2.1); **Supplementary Table 2**). The reduction in second-infection clearance time also resulted in greater symmetry between proliferation and clearance times (ratio of proliferation time to clearance time: 0.7 (0.6, 0.8) for the *n* = 1,241 BA.1/BA.2 infections with no prior infection history *vs*. 0.9 (0.7, 1.0) for the *n* = 159 BA.1/BA.2 infections with prior infection history).

### In individuals with multiple infections, second infections were cleared faster than first infections, especially when there was an intervening vaccine dose

We found no significant difference in proliferation time or peak viral concentration between first infections caused by Alpha, Delta, or other/unspecified lineages in individuals with a later Omicron BA.1/BA.2 infection (*n* = 78) vs. second infections caused by Omicron BA.1 or BA.2 in individuals with a previous Alpha, Delta, or other/unspecified infection (*n* = 157) (**Supplementary Table 1, group 2**). However, in this group, clearance times were substantially faster in second infections (5.1 days (4.7, 5.7) *vs*. 8.8 days (7.8, 9.8) in first infections, **Supplementary Table 3**). Clearance rates were also substantially faster in second infections within this group (2.4 Ct/day (2.1, 2.7) vs. 1.6 Ct/day (1.3, 1.8) in first infections, **Supplementary Table 3**). Individuals in this group who had an intervening vaccine dose between their first and second infection had a somewhat faster mean second-infection viral clearance time than those without an intervening dose (5.2 days (4.5, 9.5) with an intervening dose; 6.2 days (4.2, 8.8) without an intervening dose), though these estimates carry substantial uncertainty (**Figure 2C–F, Supplementary Table 3**). As before, the reduction in second-infection clearance time also resulted in greater symmetry between proliferation and clearance times (ratio of proliferation time to clearance time: 0.4 (0.3, 0.6) for the *n* = 78 first infections *vs*. 0.9 (0.7, 1.1) for the *n* = 157 second infections).

**Figure 2.**
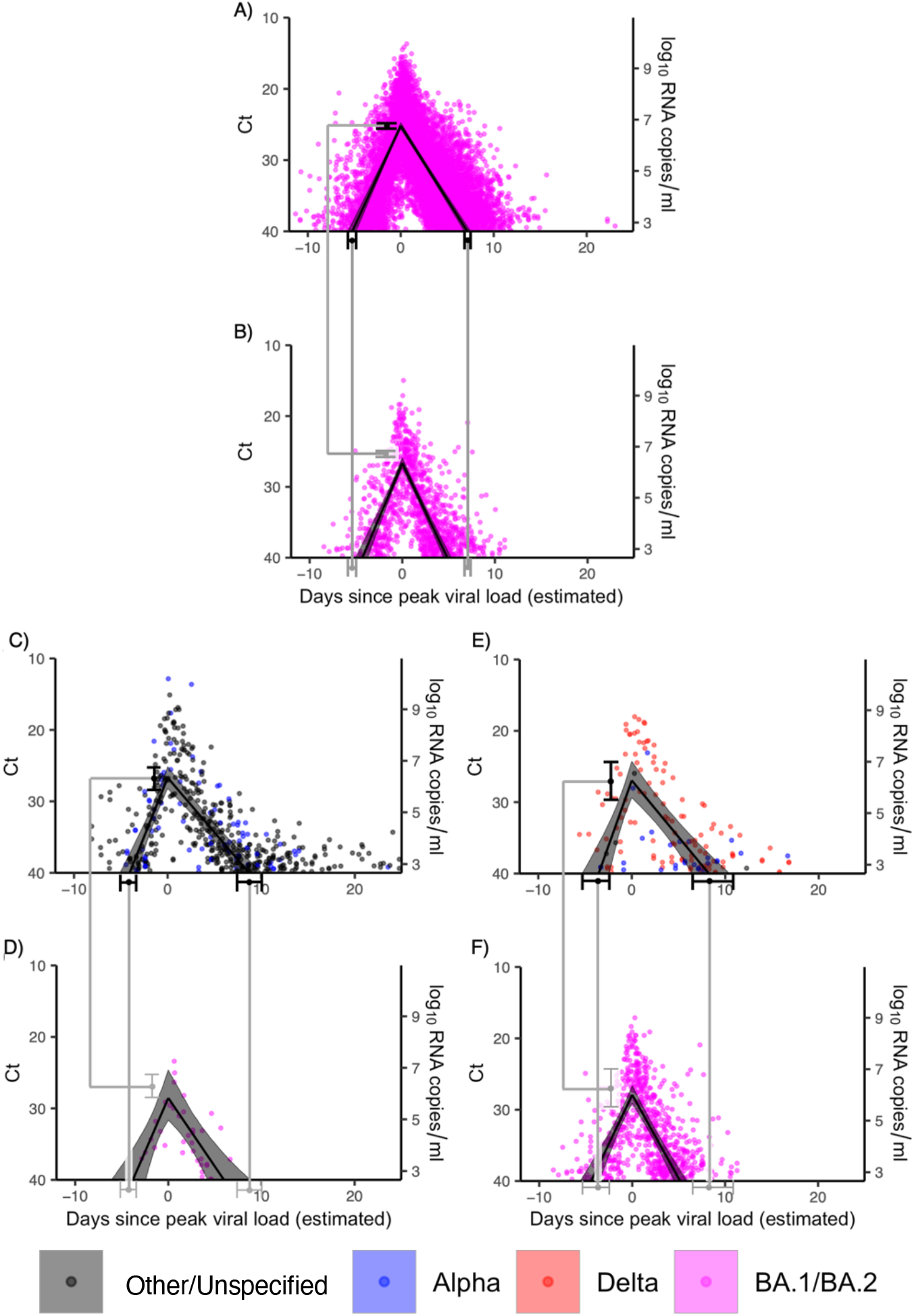
Viral kinetics of first *vs*. second infections. (A–B): Mean posterior viral trajectory (solid lines) with 95% credible interval (shaded region) for well-documented BA.1/BA.2 infections in individuals where the infection was (A) their first recorded SARS-CoV-2 infection (*n* = 1,241) or (B) their second recorded SARS-CoV-2 infection (*n* = 159; see **Supplementary Table 1, group 1**). (C–F): Mean posterior viral trajectory (solid lines) with 95% credible interval (shaded region) for well-documented infections in individuals who were infected twice during the study period. Panels (C, E) depict the viral kinetics of first Alpha/Delta/Other/Unspecified infections in unvaccinated (C, *n =* 42) and vaccinated (E, *n =* 15) individuals who later had a BA.1/BA.2 infection. Panels (D, F) depict the viral kinetics of second BA.1/BA.2 infections in individuals who did not have a vaccine dose between their first and second infection (D, *n =* 5) and in individuals who did have a vaccine dose between their first and second infection (F, *n =* 102) as well as a previous Alpha/Delta/Other/Unspecified infection (**Supplementary Table 1, group 2**, omitting any individuals with unknown vaccination status). In all panels, grey points depict the measured viral concentration for a single test. For each person, the points were shifted horizontally so that the individual’s mean posterior peak viral concentration sits at day 0. Black points and whiskers (A, C, E) depict the mean and 95% credible interval for the proliferation time, peak viral concentration, and clearance time, from left to right, for first infections. These values are repeated in grey on the lower plots (B, D, F) to facilitate comparison with the viral kinetics of second infections.

### No evidence that the kinetics of a second infection by BA.1/BA.2 differ according to the first infection’s lineage

We found no evidence the variant category of a previous infection (Alpha vs. Delta vs. other/unspecified) affected the mean viral kinetics of a later Omicron BA.1/BA.2 infection (*n* = 157, **Supplementary Table 1 group 2; Supplementary Table 5**). That is, regardless of the lineage of the initial infection, the viral kinetics of second BA.1/BA.2 infections had similar features to one another, though not to the first infection.

### An individual’s relative clearance speed is preserved across infections

Among individuals with two well-documented infections (*n* = 58; **Supplementary Table 1, group 3**), the ranking of clearance times was roughly conserved across first and second infections; that is, those with a relatively short clearance time in their first infection tended to have a short clearance time in their second infection, and *vice versa* (**Figure 3**). The Spearman correlation coefficient between individual-level mean clearance times for first *vs*. second infections was 0.475 overall (*p =* 0.0001, permutation test). When stratified by the variant category of the first infection, the Spearman correlation between individual-level mean clearance times for first *vs*. second infections was 0.783, 0.352, and 0.396 for Alpha, Delta, and infections with other/unspecified lineages, respectively. We found somewhat weaker evidence that a person’s proliferation time rank was conserved across infections, with a Spearman correlation coefficient of 0.267 (*p =* 0.02, permutation test) for first *vs*. second infections. We did not find evidence that an individual’s peak viral concentration rank was maintained across infections. These findings were robust to the choice of correlation statistic (Pearson *vs*. Spearman; **Supplementary Table 6**).

**Figure 3.**
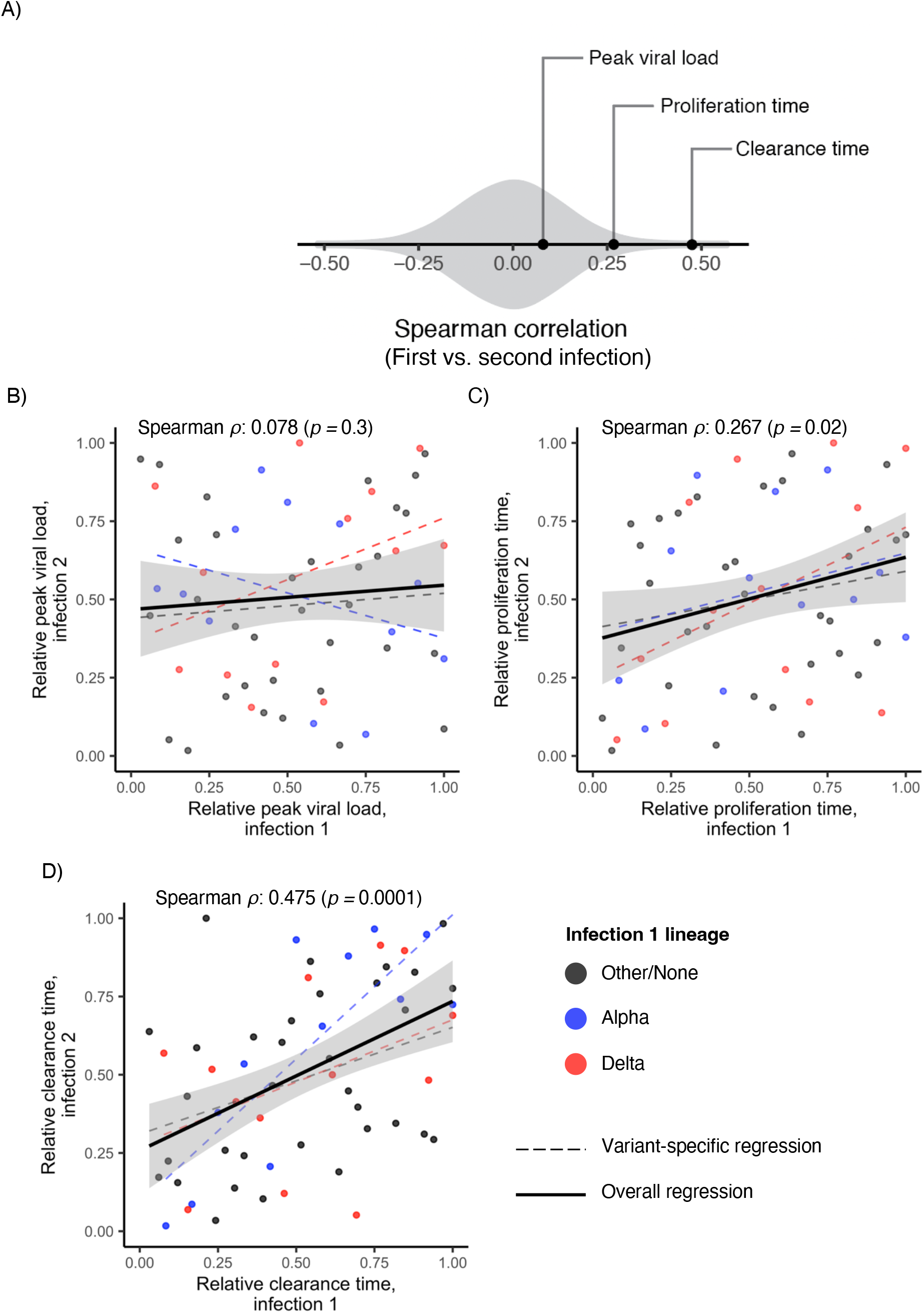
Relative viral kinetic parameters for first *vs*. second SARS-CoV-2 infections. (A): Spearman correlation (points) of peak viral load, proliferation time, and clearance time for first *vs*. second infections among *n* = 58 individuals with two well-documented infections (**Supplementary Table 1, group 3**). The height of the grey shaded area is proportional to the probability that the Spearman correlation would fall at a particular value given no correlation between first- and second-infection parameters. (B-D): Scatter plots of the normalized order statistics (points) for the peak viral concentration (B), proliferation time (C), and clearance time (D) for first *vs*. second infections in the *n =* 58 individuals with two well-documented infections. Points are colored according to the variant of the first infection (Other/None: black; Alpha: blue; Delta: red); all second infections were caused by BA.1/BA.2. Dashed lines depict the best-fit linear regression to the normalized order statistics when stratified by the variant of the first infection. The solid black line depicts the overall best-fit linear regression to the normalized order statistics combined across all first-infection variants. The grey regions depict the 95% confidence regions for this overall trend line. Order statistics were calculated by ranking the mean posterior values for each parameter (peak viral load, proliferation time, clearance time) for each infection within a variant/cardinality group (*e*.*g*., Delta/first infection). Then, these ranks were divided by the maximum rank for that variant/cardinality group, so that the normalized order statistics fall between 0 and 1.

## Discussion

In individuals with multiple infections, second infections were cleared more quickly than first infections, especially when there was an intervening vaccine dose between the two infections. The lineage of the first infection did not meaningfully affect the viral kinetics of a later BA.1/BA.2 infection. However, one’s relative speed of clearing infection persisted across infections: those with a relatively fast clearance speed in their first infection tended to have a relatively fast clearance speed in their second infection, and *vice versa*, when compared to others infected with the same variant. Thus, while prior infection and vaccination can modulate a person’s viral kinetics in absolute terms, there may also exist some further immunological mechanism, conserved across sequential infections, that determines one’s strength of immune response against SARS-CoV-2 relative to others in the population.

The mechanism underlying this persistence in clearance speed rank across subsequent infections is unclear. Some possibilities include the recency of exposure to a different related coronavirus (e.g., HKU1 or OC43),^15^ immune imprinting from early-lifetime exposure to certain coronavirus lineages,^16^ or an inherent, genome-mediated aspect of immune response. It is also unclear whether one’s relative ability to clear SARS-CoV-2 infection generalizes to other coronaviruses or to other pathogens. Serological studies and genome-wide association studies may help to illuminate the mechanisms behind persistence in SARS-CoV-2 clearance time. Such studies would be valuable for improving our basic understanding of immune response to respiratory pathogens and for developing personalized clinical respiratory disease management protocols.

A consistent finding between this and other studies on SARS-CoV-2 viral kinetics is that prior antigenic exposure, through infection or vaccination, tends to speed up viral clearance, and thus to reduce the duration of test positivity.^5,11,17^ The duration of viral positivity has various consequences both for clinical management and for public health surveillance. For clinical management, test results should be interpreted in the context of a patient’s immune history, which can modulate both the extent and expected duration of viral shedding.^5,14^ It may also be possible to adjust the recommended duration of post-infection isolation based on infection history. When estimating epidemic growth rates using cross-sectional RT-qPCR test results, it is critical to account for immune-mediated shifts in the asymmetry between viral clearance to viral proliferation times, since this asymmetry is a key component in determining whether an epidemic is growing or shrinking.^7^ We find that the difference between viral proliferation and viral clearance times decreases in second infections, which may reduce certainty in epidemic growth rates derived from cross-sectional RT-qPCR-based methods.

This study is limited by various factors. The cohort is predominately young, male, and healthy. While we adjusted for age, comorbidities and other underlying health factors were not measured. We were unable to fully disentangle the effect of both variant and first *vs*. second infection simultaneously due to small sample sizes when the population was separated into such narrow categories. Still, our findings are consistent with other studies showing that immunity tends to reduce clearance time;^5,11,17^ thus, we expect that our findings of relatively lower peak viral load and faster clearance in second infections are at least partially features of immune response and not just to viral lineage alone. This study focuses primarily on individuals who were ultimately infected twice, and these individuals may differ in important immunological and behavioral ways from those who only underwent one infection during the study period. This underscores the need for further studies that capture viral and serological kinetics in tandem.

In conclusion, immunity from a first SARS-CoV-2 infection affects the viral kinetics of a second SARS-CoV-2 infection principally by speeding up viral clearance and thus shortening the overall time of acute infection. The kinetics of a second BA.1/BA.2 infection are unaffected by the lineage of the first infection. Individuals who quickly cleared their first infection also tended to quickly clear their second infection, pointing towards persistence of underlying immune response across multiple infections. These findings help guide the interpretation of quantitative SARS-CoV-2 tests both clinically and for surveillance and point towards persistent individual-level immune mechanisms against SARS-CoV-2 that so far remain unexplained.

## Supplementary Methods

### Genome sequencing and lineage alignment

Following previously described methods, RNA was extracted from clinical samples and confirmed as SARS-CoV-2 positive.^18^ Sequencing was performed using the Illumina COVIDSeq ARTIC viral amplification primer set (V4, 384 samples, cat# 20065135). Library preparation was performed using the amplicon-based Illumina COVIDseq Test v033 and sequenced 2×74 on Illumina NextSeq 550 following Illumina’s documentation. The resulting FASTQ files were processed and analyzed on Illumina BaseSpace Labs using the Illumina DRAGEN COVID Lineage Application;^19^ versions included were 3.5.0, 3.5.1, 3.5.2, 3.5.3, and 3.5.4. The DRAGEN COVID Lineage pipeline was run with default parameters as recommended by Illumina. Lineage assignment and phylogenetics analysis were accomplished using the most recent versions of Pangolin^20^ and NextClade,^21^ respectively.

### Assessing persistence in viral kinetic parameters across infections

An individual’s viral kinetics are expected to change substantially across subsequent infections due to the accumulation of immunity through both infection and vaccination and due to differences in viral lineage, among other factors. Furthermore, the impact that these factors have on viral kinetics may be non-linear and individual-specific. Because of this, a naï ve measurement of similarity between viral kinetic parameter values in first *vs*. second infections is unlikely to yield valuable insights. However, more information might be obtained by comparing an individual’s viral kinetics against those of similar individuals, in terms of variant and immune history, in first *vs*. second infections. This is the motivation for using the Spearman correlation for assessing associations in viral kinetic parameters across subsequent infections.

To measure the Spearman correlation between first and second infections for a given viral kinetic parameter, we first assigned a rank to the mean posterior value for that parameter for all individuals in an infection cardinality/variant group. That is, within each group of (a) first infections caused by Alpha, (b) first infections caused by Delta, (c) first infections caused by other/unspecified lineages, and (d) first infections caused by BA.1/BA.2, we ranked the viral kinetic parameter (say, peak viral load) from least to greatest. Next, we divided the ranks by the maximum rank for each group, so that the normalized ranks all fell between 0 and 1. Finally, we measured the correlation between the normalized ranks for all second *vs*. first all infections; this is the Spearman correlation. We normalized the values so that all the cardinality/variant groups would have a similar scale; the Spearman correlation is unaffected by such multiplicative normalizations. As a result, individuals with a normalized order statistic close to 1 had a large estimated value for the viral kinetic parameter relative to others in their cardinality/variant group, while individuals with a normalized order statistic close to 0 had a small estimated value for the viral kinetic parameter relative to others in their cardinality/variant group.

To assess robustness to this approach, we also measured the Spearman correlations fully stratified by the variant category of the first infection. That is, for each viral kinetic parameter, we measured the Spearman correlation three times, once for individuals whose first infection was caused by Alpha, once for individuals whose first infection was caused by Delta, and one for individuals whose first infection was caused by an other/unspecified lineage. Then, when normalizing the BA.1/BA.2 order statistics, we maintained these groups, so that the second infection for an individuals who was previously infected by Alpha was only compared against other individuals who were also previously infected by Alpha. This stands in contrast to the previous approach, for which (a) all BA.1/BA.2 infections were ranked against one another regardless of the lineage of the first infection and (b) once the order statistics were calculated for each infection cardinality/variant group, all first infections were combined to generate a single regression. These values are reported in **Supplementary Table 7**.

We further assessed the sensitivity of this approach by calculating the standard Pearson correlation between the viral kinetic parameters, using the raw estimated mean posterior values for each person in their first and second infections (**Supplementary Figure 1, Supplementary Table 6**). As expected, these relationships appear to be more subject to biases due to outlying trajectories, but the general trends are the same: those with a relatively long clearance time in their first infection tended to have a long clearance time in their second infection, and *vice versa*, as measured by a positive Pearson correlation between clearance times in first *vs*. second infections.

## Data Availability

Code and data are available online at https://github.com/skissler/Ct_SequentialInfections

---

**Supplementary Table 1.**
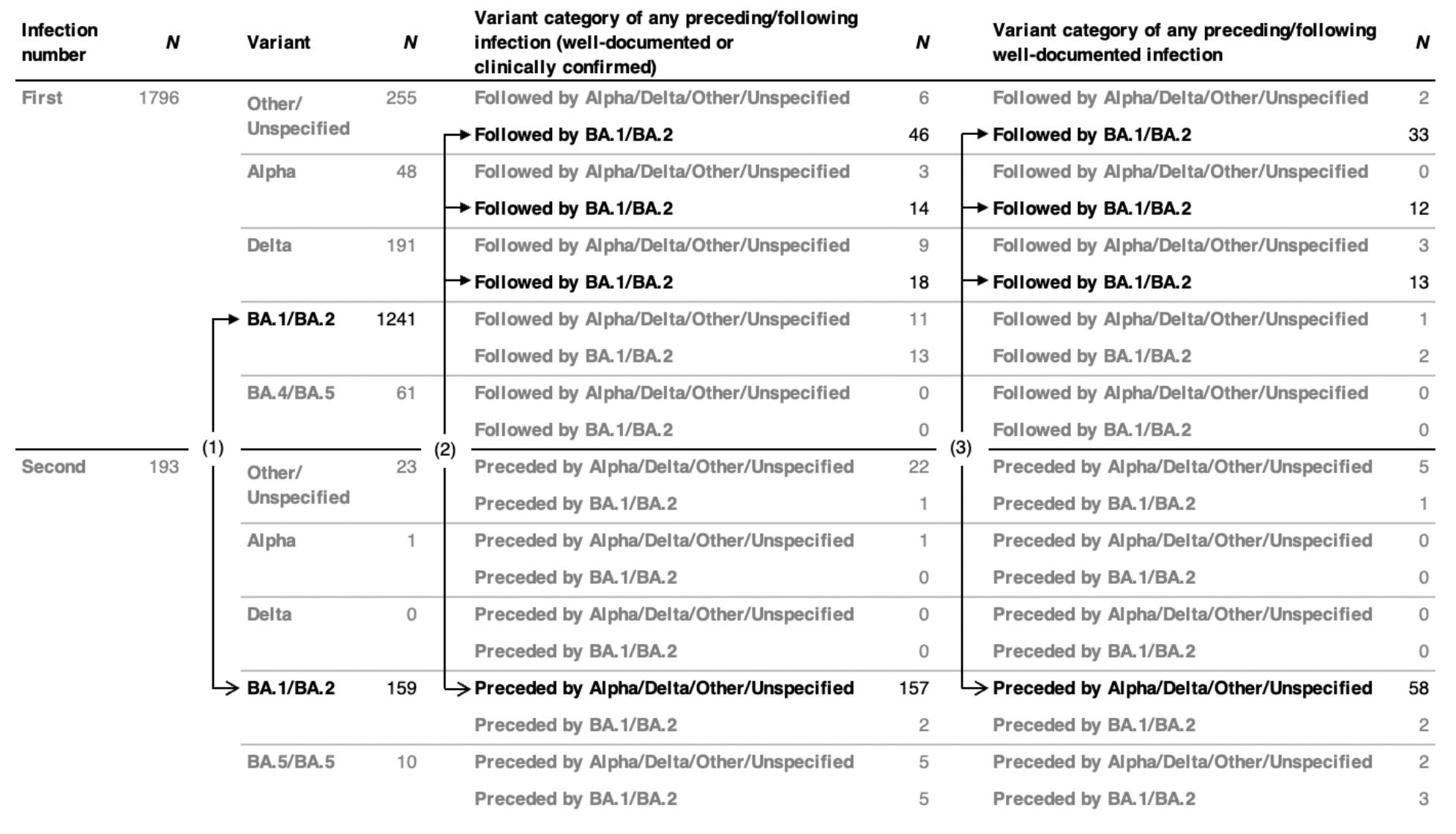
Summary of well-documented infections by variant and lineage of any preceding/following infection. Counts of well-documented infections by infection cardinality (first vs. second), variant category (Alpha, Delta, BA./BA.2, BA.4/BA.5, Other/Unspecified), and the variant category of any preceding or following infection. The main results in this manuscript reflect a comparison between (1) BA.1/BA.2 first infections (*n* = 1241) vs. BA.1/BA.2 second infections (*n* = 159), (2) Alpha/Delta/Other/Unspecified first infections that were followed by any BA.1/BA.2 infection (*n* = 78) vs. BA.1/BA.2 infections that were preceded by any Alpha/Delta/Other/Unspecified infection (*n* = 157), and (3) Alpha/Delta/Other/Unspecified first infections that were followed by a well-documented BA.1/BA.2 infection (*n* = 58) vs. BA.1/BA.2 infections that were preceded by a well-documented Alpha/Delta/Other/Unspecified infection (*n* = 58). Darker text indicates groups that are compared directly in the analyses described in the main results. Arrows connect the groups that are compared, with filled-in arrowheads denoting the “reference group” (first infections) and open arrowheads denoting the “comparison group” (second infections).

**Supplementary Table 2.** Viral kinetic parameters for BA.1/BA.2 infections, stratified by infection cardinality (first *vs*. second) and booster status and adjusted by age group.

| **Viral kinetic parameter** | **Category** | **Value** |
| --- | --- | --- |
| Peak viral concentration (Ct) | First infection, unboosted | 25.8 (24.8, 26.7) |
|  | First infection, boosted | 25.9 (25.3, 26.5) |
|  | Second infection, unboosted | 26.7 (25.0, 28.2) |
|  | Second infection, boosted | 27.0 (25.9, 28.0) |
| Peak viral concentration (GE/ml) | First infection, unboosted | 6.6 (6.3, 6.9) |
|  | First infection, boosted | 6.6 (6.4, 6.7) |
|  | Second infection, unboosted | 6.4 (5.9, 6.8) |
|  | Second infection, boosted | 6.3 (6.0, 6.6) |
| Proliferation time (days) | First infection, unboosted | 4.4 (3.8, 5.2) |
|  | First infection, boosted | 5.2 (4.7, 5.9) |
|  | Second infection, unboosted | 3.6 (2.7, 4.7) |
|  | Second infection, boosted | 4.6 (3.7, 5.7) |
| Clearance time (days) | First infection, unboosted | 6.0 (5.5, 6.6) |
|  | First infection, boosted | 7.1 (6.7, 7.6) |
|  | Second infection, unboosted | 4.5 (3.8, 5.3) |
|  | Second infection, boosted | 5.1 (4.5, 5.7) |
| Proliferation rate (Ct/day) | First infection, unboosted | 3.2 (2.7, 3.8) |
|  | First infection, boosted | 2.7 (2.4, 3.1) |
|  | Second infection, unboosted | 3.8 (2.7, 5.1) |
|  | Second infection, boosted | 2.9 (2.2, 3.6) |
| Clearance rate (Ct/day) | First infection, unboosted | 2.4 (2.1, 2.6) |
|  | First infection, boosted | 2.0 (1.8, 2.1) |
|  | Second infection, unboosted | 3.0 (2.4, 3.6) |
|  | Second infection, boosted | 2.6 (2.2, 3.0) |

**Supplementary Table 3.**
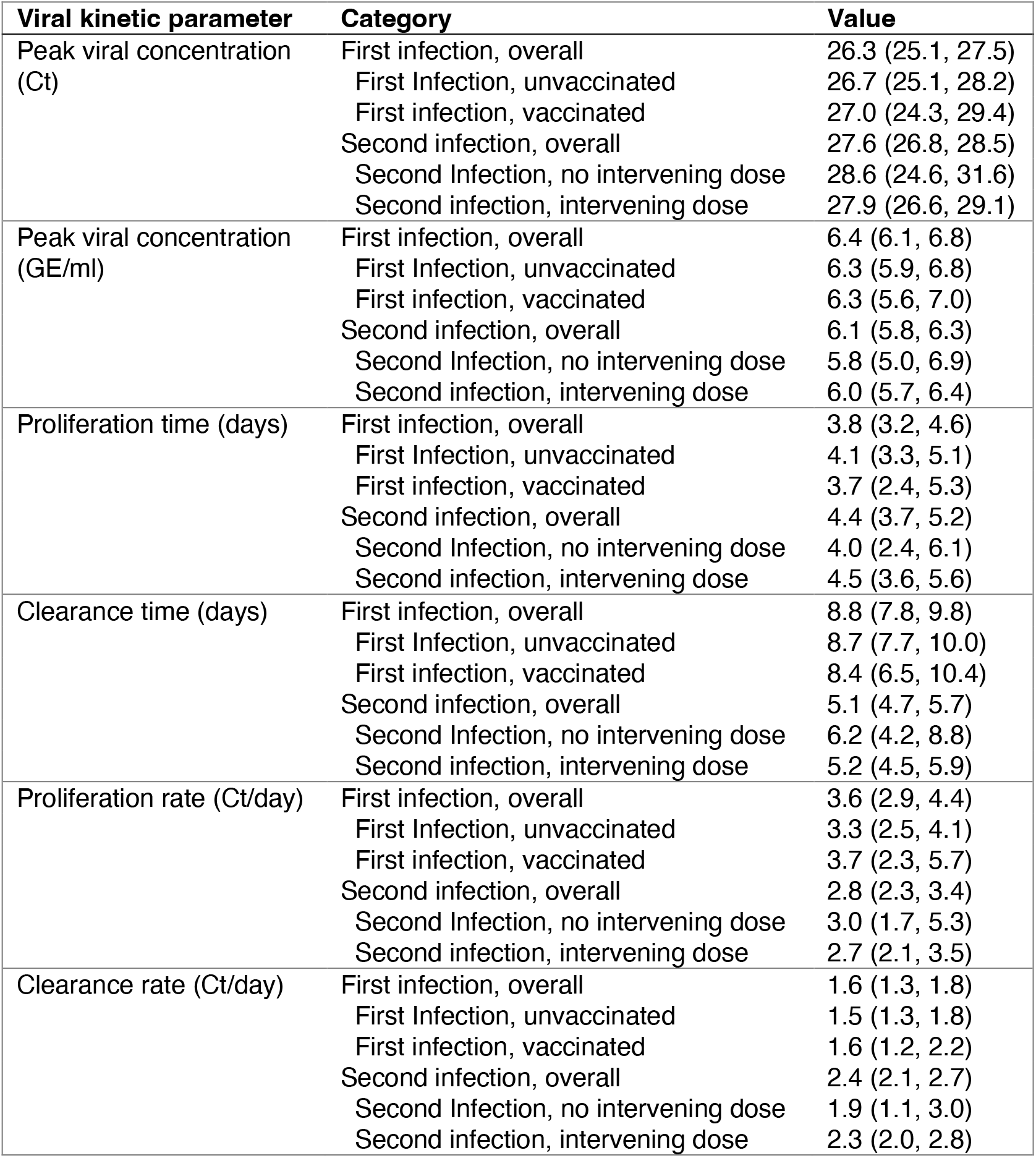
Viral kinetic parameters for first *vs*. second infections in individuals with multiple infections, stratified by vaccination status.

| **Viral kinetic parameter** | **Category** | **Value** |
| --- | --- | --- |
| Peak viral concentration (Ct) | First infection, overall | 26.3 (25.1, 27.5) |
|  | First Infection, unvaccinated | 26.7 (25.1, 28.2) |
|  | First infection, vaccinated | 27.0 (24.3, 29.4) |
|  | Second infection, overall | 27.6 (26.8, 28.5) |
|  | Second Infection, no intervening dose | 28.6 (24.6, 31.6) |
|  | Second infection, intervening dose | 27.9 (26.6, 29.1) |
| Peak viral concentration (GE/ml) | First infection, overall | 6.4 (6.1, 6.8) |
|  | First Infection, unvaccinated | 6.3 (5.9, 6.8) |
|  | First infection, vaccinated | 6.3 (5.6, 7.0) |
|  | Second infection, overall | 6.1 (5.8, 6.3) |
|  | Second Infection, no intervening dose | 5.8 (5.0, 6.9) |
|  | Second infection, intervening dose | 6.0 (5.7, 6.4) |
| Proliferation time (days) | First infection, overall | 3.8 (3.2, 4.6) |
|  | First Infection, unvaccinated | 4.1 (3.3, 5.1) |
|  | First infection, vaccinated | 3.7 (2.4, 5.3) |
|  | Second infection, overall | 4.4 (3.7, 5.2) |
|  | Second Infection, no intervening dose | 4.0 (2.4, 6.1) |
|  | Second infection, intervening dose | 4.5 (3.6, 5.6) |
| Clearance time (days) | First infection, overall | 8.8 (7.8, 9.8) |
|  | First Infection, unvaccinated | 8.7 (7.7, 10.0) |
|  | First infection, vaccinated | 8.4 (6.5, 10.4) |
|  | Second infection, overall | 5.1 (4.7, 5.7) |
|  | Second Infection, no intervening dose | 6.2 (4.2, 8.8) |
|  | Second infection, intervening dose | 5.2 (4.5, 5.9) |
| Proliferation rate (Ct/day) | First infection, overall | 3.6 (2.9, 4.4) |
|  | First Infection, unvaccinated | 3.3 (2.5, 4.1) |
|  | First infection, vaccinated | 3.7 (2.3, 5.7) |
|  | Second infection, overall | 2.8 (2.3, 3.4) |
|  | Second Infection, no intervening dose | 3.0 (1.7, 5.3) |
|  | Second infection, intervening dose | 2.7 (2.1, 3.5) |
| Clearance rate (Ct/day) | First infection, overall | 1.6 (1.3, 1.8) |
|  | First Infection, unvaccinated | 1.5 (1.3, 1.8) |
|  | First infection, vaccinated | 1.6 (1.2, 2.2) |
|  | Second infection, overall | 2.4 (2.1, 2.7) |
|  | Second Infection, no intervening dose | 1.9 (1.1, 3.0) |
|  | Second infection, intervening dose | 2.3 (2.0, 2.8) |

**Supplementary Table 4.** Viral kinetic parameters by variant.

| **Viral kinetic parameter** | **Category** | **Value** |
| --- | --- | --- |
| Peak viral concentration (Ct) | Other/None | 26.7 (25.2, 28.0) |
|  | Alpha | 27.2 (24.7, 29.5) |
|  | Delta | 24.4 (21.8, 26.7) |
|  | BA.1/BA.2 | 27.5 (26.6, 28.4) |
| Peak viral concentration (GE/ml) | Other/None | 6.3 (6.0, 6.7) |
|  | Alpha | 6.2 (5.6, 6.9) |
|  | Delta | 7.0 (6.3, 7.7) |
|  | BA.1/BA.2 | 6.1 (5.9, 6.4) |
| Proliferation time (days) | Other/None | 4.0 (3.3, 4.9) |
|  | Alpha | 3.6 (2.6, 5.0) |
|  | Delta | 3.4 (2.3, 4.6) |
|  | BA.1/BA.2 | 4.4 (3.7, 5.2) |
| Clearance time (days) | Other/None | 9.1 (8.1, 10.3) |
|  | Alpha | 9.0 (7.2, 11.2) |
|  | Delta | 7.8 (6.3, 9.5) |
|  | BA.1/BA.2 | 5.1 (4.6, 5.7) |
| Proliferation rate (Ct/day) | Other/None | 3.3 (2.6, 4.1) |
|  | Alpha | 3.6 (2.4, 5.2) |
|  | Delta | 4.8 (3.2, 7.0) |
|  | BA.1/BA.2 | 2.9 (2.3, 3.5) |
| Clearance rate (Ct/day) | Other/None | 1.5 (1.2, 1.7) |
|  | Alpha | 1.4 (1.1, 1.9) |
|  | Delta | 2.0 (1.6, 2.6) |
|  | BA.1/BA.2 | 2.4 (2.1, 2.8) |

**Supplementary Table 5.** Viral kinetic parameters for second infections in individuals with multiple infections, stratified by the variant of the first infection.

| **Viral kinetic parameter** | **Category** | **Value** |
| --- | --- | --- |
| Peak viral concentration (Ct) | Other/None | 27.3 (26.3, 28.2) |
|  | Alpha | 27.2 (25.1, 29.3) |
|  | Delta | 27.5 (25.3, 29.2) |
| Peak viral concentration (GE/ml) | Other/None | 6.2 (5.9, 6.5) |
|  | Alpha | 6.2 (5.6, 6.8) |
|  | Delta | 6.1 (5.6, 6.7) |
| Proliferation time (days) | Other/None | 4.4 (3.7, 5.2) |
|  | Alpha | 4.6 (3.3, 6.3) |
|  | Delta | 4.8 (3.5, 6.6) |
| Clearance time (days) | Other/None | 5.0 (4.4, 5.6) |
|  | Alpha | 5.2 (4.1, 6.6) |
|  | Delta | 4.9 (3.8, 6.2) |
| Proliferation rate (Ct/day) | Other/None | 2.9 (2.4, 3.5) |
|  | Alpha | 2.9 (1.9, 4.1) |
|  | Delta | 2.7 (1.8, 3.8) |
| Clearance rate (Ct/day) | Other/None | 2.6 (2.2, 2.9) |
|  | Alpha | 2.5 (1.8, 3.3) |
|  | Delta | 2.6 (1.9, 3.5) |

**Supplementary Table 6.** Correlation between viral kinetic parameters in first *vs*. second infections in individuals with two well-documented infections. Significance was calculated using a permutation test, with the *p*-value representing the proportion of draws that exceeded the calculated significance value under the assumption of no association in viral kinetic parameters between the first and second infection. Bold values indicate significance at the *p* = 0.05 level.

| Viral kinetic parameter | Correlation statistic | Value | Significance ($p$) |
| --- | --- | --- | --- |
| Peak viral concentration (Ct) | Spearman | 0.0799 | 0.278 |
|  | Pearson | 0.128 | 0.172 |
| Proliferation time (days) | Spearman | **0.267** | **0.0206** |
|  | Pearson | **0.242** | **0.0396** |
| Clearance time (days) | Spearman | **0.475** | **0.0001** |
|  | Pearson | **0.336** | **0.0081** |

**Supplementary Table 7.** Correlation between viral kinetic parameters in first *vs*. second infections in individuals with two well-documented infections, stratified by the lineage of the first infection. Significance was calculated using a permutation test, with the *p*-value representing the proportion of draws that exceeded the calculated significance value under the assumption of no association in viral kinetic parameters between the first and second infection. Bold values indicate significance at the *p* = 0.05 level.

| Viral kinetic parameter | Correlation statistic | First infection lineage | Value | Significance ($p$) |
| --- | --- | --- | --- | --- |
| Peak viral concentration (Ct) | Spearman | Alpha | -0.308 | 0.828 |
|  |  | Delta | 0.330 | 0.132 |
|  |  | Other/Unspecified | 0.0675 | 0.346 |
|  | Pearson | Alpha | -0.299 | 0.828 |
|  |  | Delta | 0.416 | 0.0717 |
|  |  | Other/Unspecified | 0.0627 | 0.357 |
| Proliferation time (days) | Spearman | Alpha | 0.245 | 0.218 |
|  |  | Delta | 0.418 | 0.074 |
|  |  | Other/Unspecified | 0.180 | 0.156 |
|  | Pearson | Alpha | 0.309 | 0.166 |
|  |  | Delta | 0.462 | 0.0857 |
|  |  | Other/Unspecified | **0.310** | **0.0241** |
| Clearance time (days) | Spearman | Alpha | **0.783** | **0.0019** |
|  |  | Delta | 0.352 | 0.114 |
|  |  | Other/Unspecified | **0.396** | **0.0121** |
|  | Pearson | Alpha | **0.684** | **0.0054** |
|  |  | Delta | **0.486** | **0.0525** |
|  |  | Other/Unspecified | 0.233 | 0.108 |

**Supplementary Figure 1.**
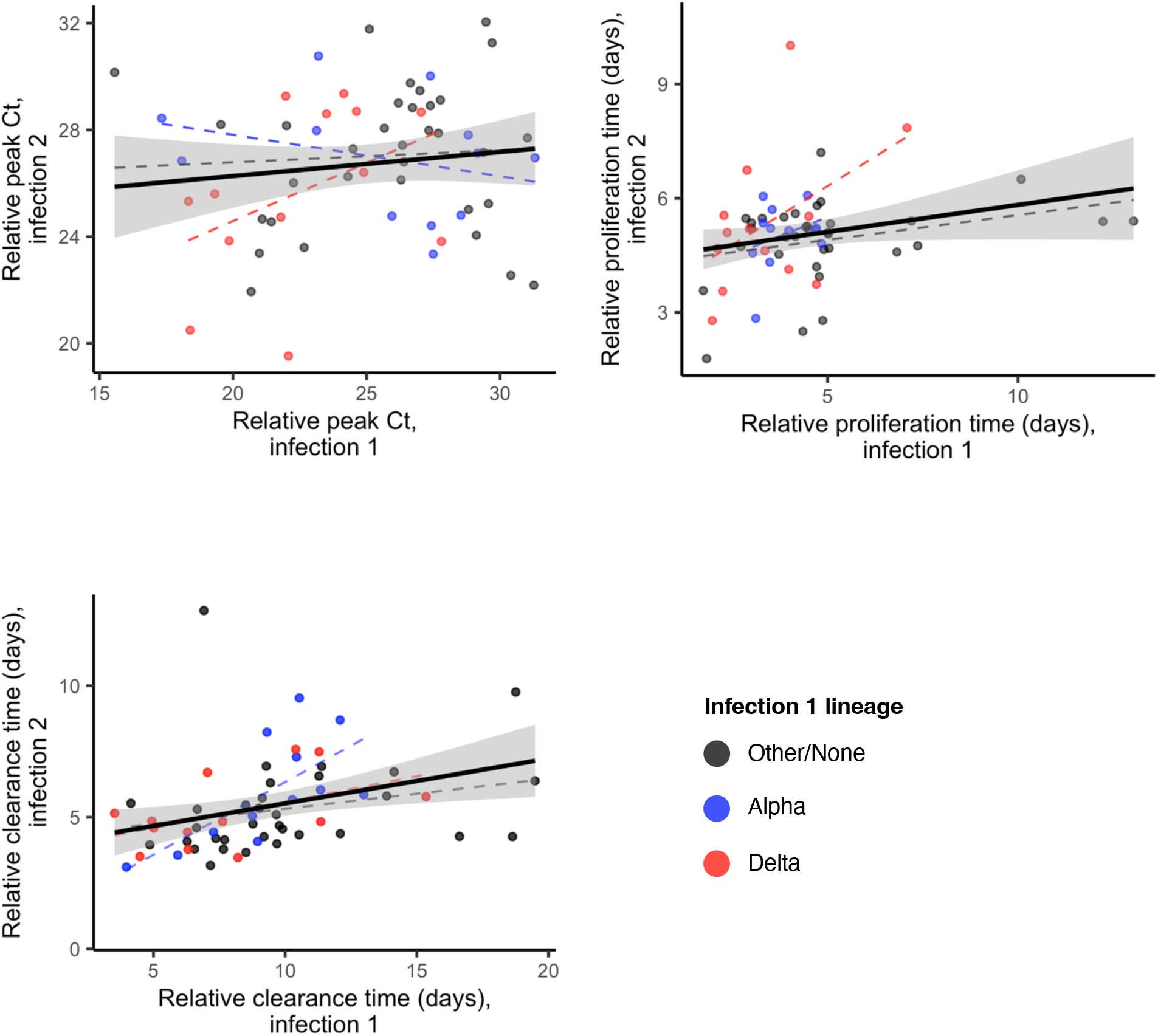
Relative viral kinetic parameters for first *vs*. second SARS-CoV-2 infections. (A-C): Scatter plots of the raw individual-level mean posterior viral kinetic parameter values (points) for the peak viral concentration (A), proliferation time (B), and clearance time (C) for first *vs*. second infections in individuals with two well-documented infections. Points are colored according to the variant of the first infection (Other/None: black; Alpha: blue; Delta: red); all second infections were caused by BA.1/BA.2. Dashed lines depict the best-fit linear regression to the parameter values when stratified by the variant of the first infection. The solid black line depicts the overall best-fit linear regression to the mean posterior parameter values combined across all first-infection variants. The grey regions depict the 95% confidence regions for this overall trend line.

## Acknowledgements

The authors acknowledge Radhika Samant and Sarah Connolly for assistance with data curation.

## Data and code availability

All data and code needed to reproduce the findings in this manuscript may be accessed in the following repository: https://github.com/skissler/Ct_SequentialInfections

